# Dynamic Biobanking for Advancing Breast Cancer Research

**DOI:** 10.1101/2022.12.15.22283484

**Authors:** Maryam Abdollahyan, Emanuela Gadaleta, Millahat Asif, Jorge Oscanoa, Rachel Barrow-McGee, Samantha Jones, Louise Jones, Claude Chelala

## Abstract

Longitudinal patient biospecimens and data advance breast cancer research through enabling precision medicine approaches for identifying risk, early diagnosis, improved disease management and targeted therapy. Cancer biobanks must evolve to provide not only access to high-quality annotated biospecimens and rich associated data, but also the tools required to harness these data. We present the Breast Cancer Now Tissue Bank centre at the Barts Cancer Institute as an exemplar of a dynamic biobanking ecosystem that hosts and links longitudinal biospecimens and multimodal data including electronic health records, genomic and imaging data, offered alongside integrated data sharing and analytics tools. We demonstrate how such an ecosystem can inform precision medicine efforts in breast cancer research.

**Summary:** We present the infrastructure established for the Breast Cancer Now Tissue Bank centre at the Barts Cancer Institute as an exemplar of a dynamic biobank that supports precision medicine for breast cancer through providing access to integrated data and tools, and could serve as a blueprint for future cancer biobanks.

## 1. Introduction

Approximately 1 in 8 women are diagnosed with breast cancer in their lifetime [1]. Despite recent advances in early detection and treatment of BC, and extensive histopathological and molecular characterisation of primary tumours, around 30% of patients go on to experience recurrence [2] or metastasis which is considered incurable. These figures are expected to rise in the upcoming years due to the disruptions in screening programmes and treatments related to the COVID-19 pandemic, with nearly 1 million women in the UK having missed their mammogram appointments [3]. While effects of the COVID-19 pandemic amplified existing challenges and health inequalities, increasing demand for innovative solutions, the risk factors for breast cancer remain not well-understood. This is particularly evident in multiethnic communities who are underrepresented in or absent from large studies. Cancer biobanks are important biomedical research infrastructures for preclinical, translational and clinical studies that can help address these challenges.

As we move forward in the era of precision medicine, cancer biobanks are required to adopt new strategies for patient recruitment and collection, processing, storage and distribution of biospecimens and data to better contribute to this research area and the wider healthcare landscape. Linkage to other data sources such as electronic health records (EHRs) is vital to complement a biobank’s clinical data. Healthcare datasets, as managed in EHR systems, are typically stored in highly variable formats. There is homogeneity in terms of structure and adoption of standard biomedical ontologies (e.g., ICD [4], OPCS [5] and SNOMED-CT [6]); however, a notable portion of EHRs (e.g., discharge summaries) is in a free-text format. While these unstructured data are often used to report the results of examinations, tests or procedures, and serve as a means of communication between healthcare providers, they contain valuable information that is not possible to obtain from their structured counterparts. Such information can be extracted using artificial intelligence (AI) algorithms and stored in a structured format for subsequent processing. Similarly, when combined with a biobank’s clinical data, sequencing data allow researchers to discover new variants that contribute to disease onset and progression, identify drug targets for accelerating drug discovery and development, and stratify patients into groups such as those more likely to respond to treatment or experience side-effects. Moreover, provision of tools for analysing and sharing these data is crucial to maximise the use of the biospecimen and data hosted by cancer biobanks.

Breast Cancer Now Tissue Bank (BCNTB) [7] is the UK’s first national breast cancer biobank, providing access to the largest collection of longitudinal biospecimens and data from breast cancer patients and individuals without the disease at various stages of the care pathway. The Bank has 8 (4 legacy and 4 active) centres in England and Scotland, with Barts Cancer Institute (BCI) acting as the coordinating centre. In this paper, we explore the biospecimens, data and tools made available by the BCNTB centre at BCI (BCNTB-BCI). We present an approach to building and maintaining a dynamic biobanking ecosystem, that features: A centralised database with built-in quality assurance mechanisms for storing multimodal data; linkage to data from external sources, including EHRs from primary and secondary care plus genomic data from national and international sequencing initiatives, for enriching the Bank’s data; and an integrated biospecimen and data request system and a bioinformatics portal for sharing and analysing these data. While individual elements of such an infrastructure have been suggested in the past (e.g., see [8–10]), no cancer biobank has implemented all these elements, specifically for breast cancer research. Using BCNTB-BCI as an exemplar, we demonstrate how dynamic cancer biobanks can fill the gaps in breast cancer patient data, support researchers to advance breast cancer research and assist clinicians in early diagnosis and effective treatment of breast cancer for optimal outcome.

## 2. Materials and Methods

### Multimodal Data

To date, BCNTB-BCI has collected biospecimens and data from over 2,400 breast cancer patients and individuals without the disease including those who have had cosmetic breast procedures or are at high risk of BC. These biospecimens include liquid samples (e.g., whole blood, plasma, serum and buffy coat), fresh and frozen tissue samples (e.g., tissue microarrays (TMAs), formalin-fixed paraffin-embedded (FFPE) blocks and Haemotoxylin and Eosin (H&E) stains) and primary cells isolated from breast tissue (e.g., epithelial and myoepithelial cells and fibroblasts). The associated data are manually curated by the Bank through a variety of sources (e.g., patient questionnaires and the East London Patient Record (eLPR) system [11]). These data include donor demographics and survival status, personal and family medical history, lifestyle, presentation and diagnosis, procedures (biopsy, surgery and imaging), treatments (chemotherapy and radiotherapy), follow-up reports and images (scanned H&E slides). As a result, BCNTB-BCI hosts a repository of multimodal clinical, biospecimen and imaging data.

Data from BCNTB-BCI and all other BCNTB centres are stored in a centralised Research Electronic Data Capture (REDCap) [12,13] database, a secure web-based data capture application for building and managing online surveys and databases to support research studies, at the Bank’s coordinating centre. Several REDCap features are utilised to enable and improve multisite access, data entry and validation (via branching logic, calculated fields and custom scripts run through the REDCap API), audits (e.g., via custom reports) and regulatory compliance. Furthermore, BCNTB-BCI data are mapped to concepts from the SNOMED-CT biomedical ontology to facilitates the exchange and comparison of these data with information from other sources (e.g., EHRs and genomic data).

### Linkage to EHRs and Genomic Data

To supplement and validate the Bank’s manually curated internal data, we considered linkage to external data from primary and secondary care as well as national and international sequencing initiatives. BCNTB-BCI data are linked to EHRs from Barts Health NHS Trust (BH) via the National Health Service (NHS) or hospital number. BH is the largest NHS trust in London with 5 hospitals (St Bartholomewʹs, The Royal London, Mile End, Newham and Whipps Cross) serving 2.5 million people across East London [14]. These EHRs include patient demographics and survival status, medical history, physical and social assessments, diagnosis, procedures, imaging reports, pathology results and histopathology reports, electronic prescriptions, maternity services data, clinician notes and cancer registries data (e.g., the Somerset Cancer Register [15]) that are consolidated from several datasets hosted or linked by BH [16]. To transform EHRs from BH into clinically relevant features for construction of cohorts suitable for breast cancer research, we performed a series of data cleaning and wrangling processes. In addition, we used Natural Language Processing (NLP) techniques to extract a variety of information, including symptoms and outcomes, from the unstructured part of these EHRs, and stored it in a structured format (e.g., mapped to the SNOMED-CT concepts). For instance, we applied NLP to over 45,000 free-text reports of various imaging procedures (e.g., mammogram, computed tomography (CT), magnetic resonance imaging (MRI) and ultrasound (US)) to extract information that can be used to predict breast cancer recurrence (Abdollahyan et al., manuscript in preparation).

To date, tumour and normal samples from many BCNTB donors have been sequenced and the results made available to researchers through repositories such as the European Genome-phenome Archive (EGA) [17]. Moreover, BCNTB-BCI data are linked to sequencing data from the 100,000 Genomes Project (100K GP) [18] via a unique ID for over 320 BCNTB donors who are dually consented by BCNTB-BCI and 100K GP. These data, where available, include primary clinical data for the 100K GP participants and secondary clinical data received from NHS Digital and the National Cancer Registration and Analysis Service [19].

Biospecimen and Data Request System. The Bank’s expression of interest (EOI) and application forms are implemented in REDCap. The forms are accompanied by Sample Finder, a tool implemented using an open-source Python web development framework [20] that allows researchers to search and filter biospecimens by donor and tumour characteristics. Data shown in Sample Finder are exported from the centralised REDCap database where BCNTB data are stored, reflecting the most up-to-date clinical and sample data available at the time. Using Sample Finder, researchers can instantly see how many cases match their defined cohort and view a summary report in the form of a graphical breakdown of the matching cases’ donor and tumour statistics. In case of successful biospecimen and data request applications, researchers are invited to return the data generated as part of their study to the Bank after the study results enter the public domain. The Biospecimen and Data Request System is available at https://breastcancernow.org/breastcancer-research/breast-cancer-now-tissue-bank/how-apply-tissue-bank.

Bioinformatics Portal. The Bank hosts its own bioinformatics portal, implemented using open-source Python and R packages [21], that includes Analytics Hub, a data analytics platform. The portal acts as a central hub, giving researchers access to a list of associated publications, user guides and the Bank’s contact information.

Analytics Hub, powered by a custom version of SNPnexus [22], provides an environment on which the output of Whole Genome Sequencing pipelines (e.g., FASTQ and VCF files), clinical and omics data returned by studies that used BCNTB-BCI biospecimens (e.g., the Spatial Characterisation [23] and the 100K GP cohort datasets) and publicly available data from large-scale studies (e.g., the Cancer Genome Atlas (TCGA) [24] and the Cancer Cell Line Encyclopedia (CCLE) [25] datasets) can be viewed, filtered, analysed and incorporated into research projects and grant applications. Analytics Hub has bidirectional links to Sample Finder, i.e., upon entering a query in Analytics Hub, the platform also checks the availability of biospecimens and data from BCNTB-BCI donors with the same characteristics on Sample Finder, for which researchers can submit an EOI to the Bank; inversely, upon entering a query in Sample Finder, the tool also checks the availability of data for patients from other studies with the same characteristics of interest, which researchers can explore on Analytics Hub. The Bioinformatics Portal is available at https://bcntb.bcc.qmul.ac.uk/home.

## 3. Results

### 3.1. BCNTB-BCI Infrastructure

Figure 1 shows an overview of the described BCNTB-BCI infrastructure.

**Figure 1.**
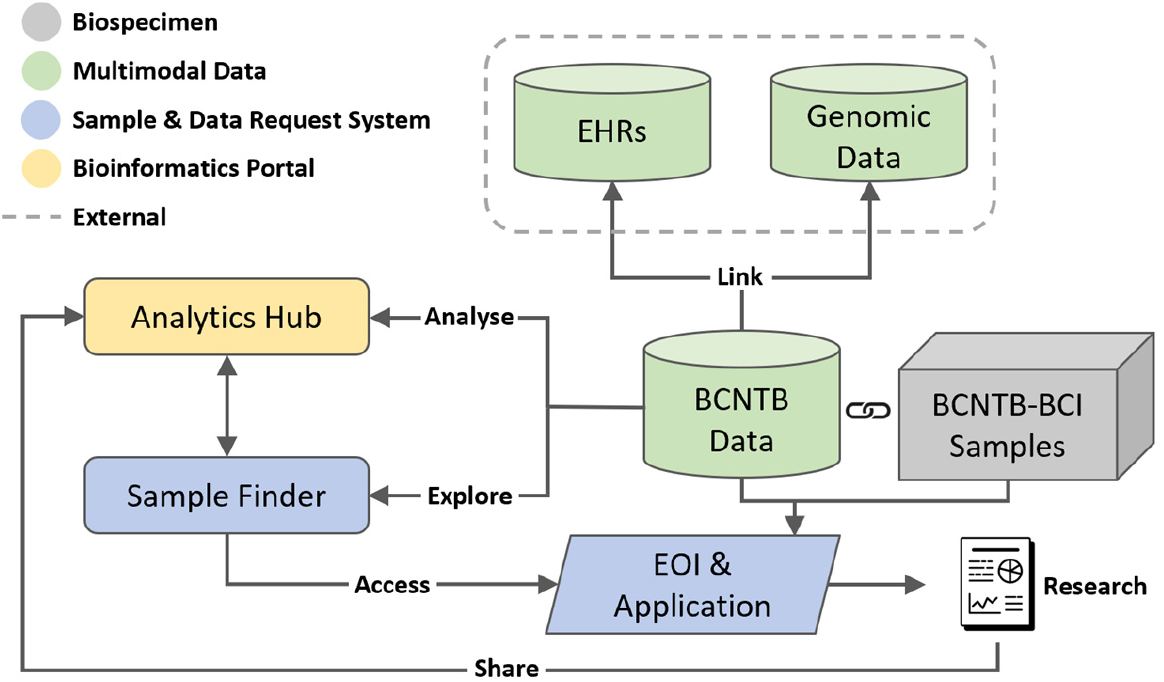
Overview of BCNTB-BCI infrastructure showing the centralised REDCap database where BCNTB data are stored, linkage to EHRs from BH and genomic data from 100K GP, the integrated Biospecimen and Data Request System and Bioinformatics Portal.

### 3.2. BCNTB-BCI Cohort Overview

Linkage of BCNTB-BCI data to EHRs allows a quick inspection of the breast cancer landscape in our population. Figures 2, 3 and 4 show the evolution of disease status in the Bank’s donors over 5 years of follow-up (the period during which risk of breast cancer recurrence is high), their age at diagnosis of primary breast cancer and ethnicity, and the cooccurrence of conditions that are often comorbid with breast cancer in these patients, respectively. Our results indicate that breast cancer screening uptake in the population of East London covered by BH is low, resulting in many cases presenting at later stages of the disease. For instance, the de novo metastatic breast cancer rate is at least 2% and the incidence of recurrence is highest in the first year after diagnosis (Figure 2). This is in keeping with the national statistics. Additionally, the results show that the most common comorbid condition is cardiovascular disease with a prevalence rate of 38%, followed by hypertension and rheumatic disease (Figure 4). This is in agreement with statistics reported in numerous other studies that investigated the prevalence of comorbidities in breast cancer patients (e.g., see [26]). However, our results also highlight the challenges in breast cancer detection and treatment in the multi-ethnic population of East London, where over 20% and over 30% of patients are Black and Asian [14], respectively, with survival being worse in these ethnic groups. For example, the 5-year survival rate is 75%, which is significantly lower than the national average of 85% [1] (Figure 2), and the age at diagnosis is younger, particularly in the Asian and Black groups (Figure 3). In addition, the results highlight the challenges in breast cancer management in this region. For example, the comorbidity burden is high, with 14% of patients experiencing 3 or more comorbidities (Figure 4).

**Figure 2.**
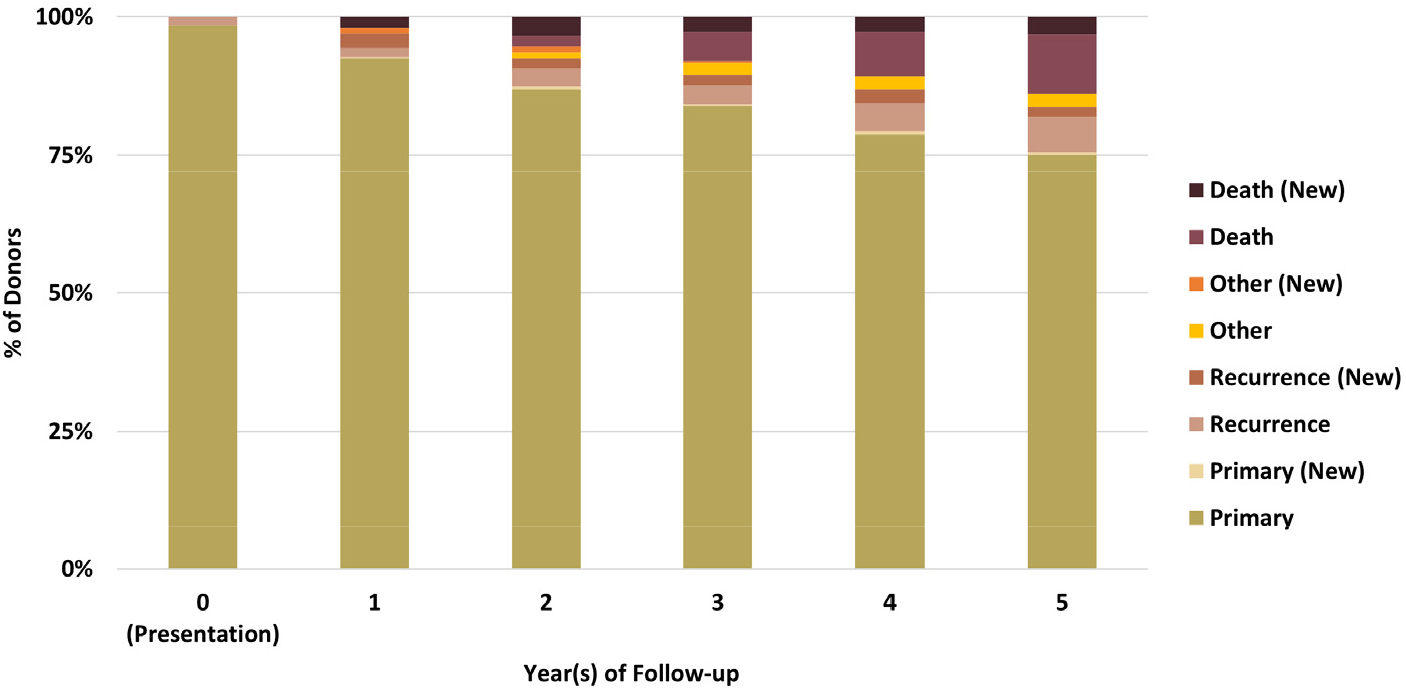
Evolution of disease status in BCNTB-BCI donors over 5 years of follow-up (the ‘Other’ category consists of cosmetic and high-risk cases; new and existing cases at the end of each follow-up year are shown separately).

**Figure 3.**
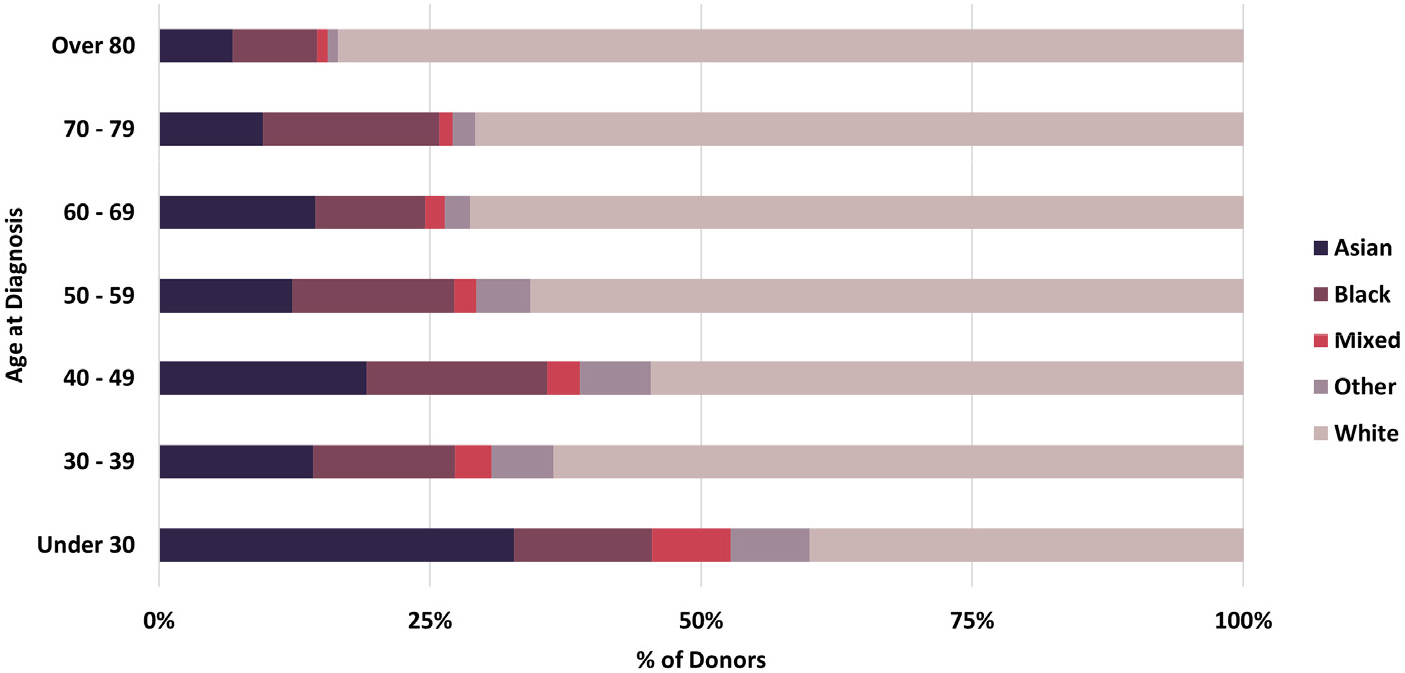
Age at diagnosis of primary breast cancer and ethnicity of BCNTB-BCI patients.

**Figure 4.**
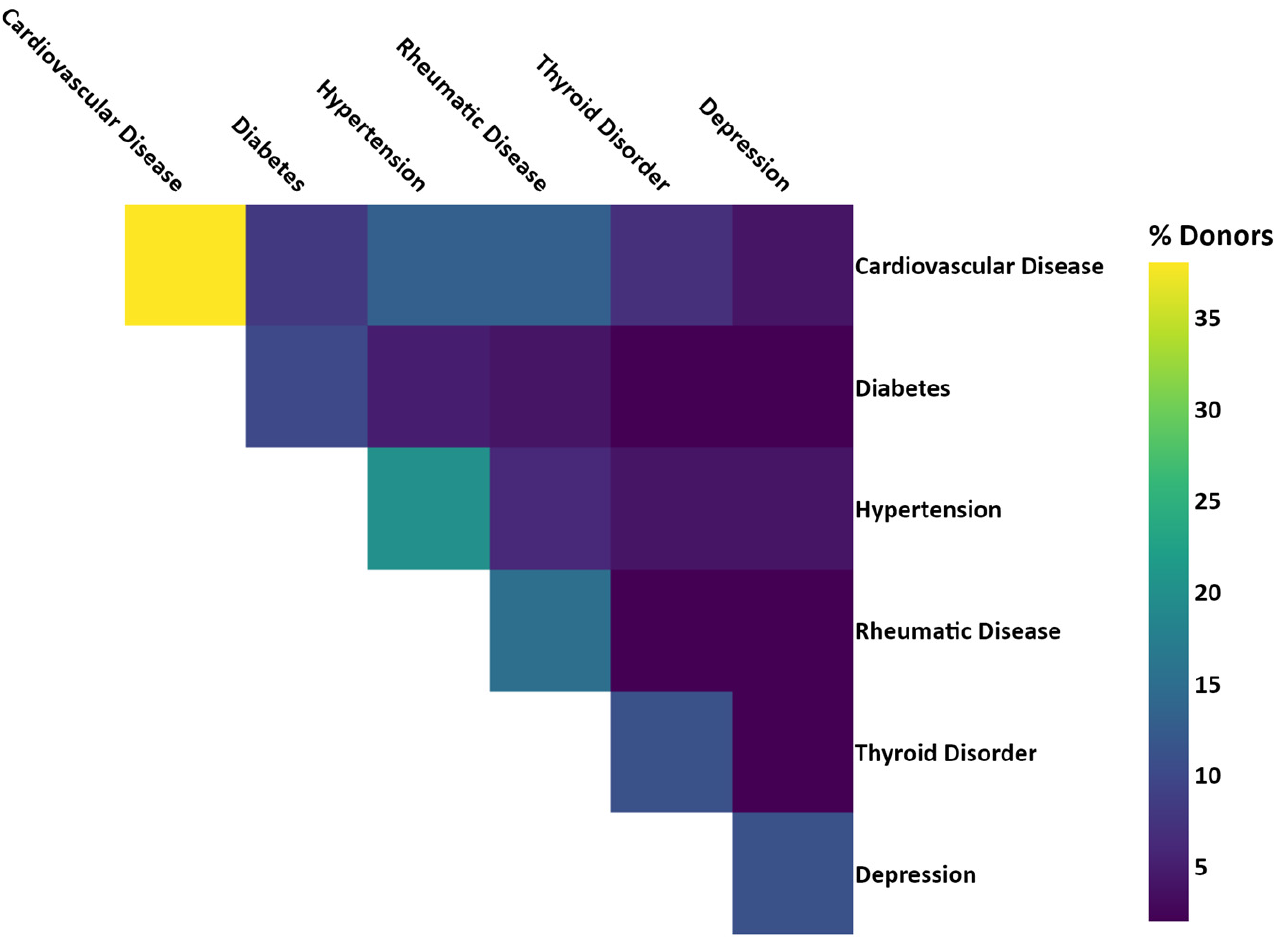
Co-occurrence of comorbid conditions in BCNTB-BCI patients (the numbers are calculated based on the ICD-10 codes used to compute the Elixhauser comorbidity index [27]).

### 3.3. Clinical and Molecular Patient Journey Narratives

The infrastructure of our cancer biobank facilitates the generation of clinical and molecular patient journey narratives, which are powerful tools for precision medicine. Figure 5 shows the clinical patient journey narrative for a breast cancer patient over 5 years (denoted by I to V in the figures) based on their biobank data from BCNTB-BCI. These data are mostly structured, which eases visualisation and interpretation of patient journey narratives.

**Figure 5.**
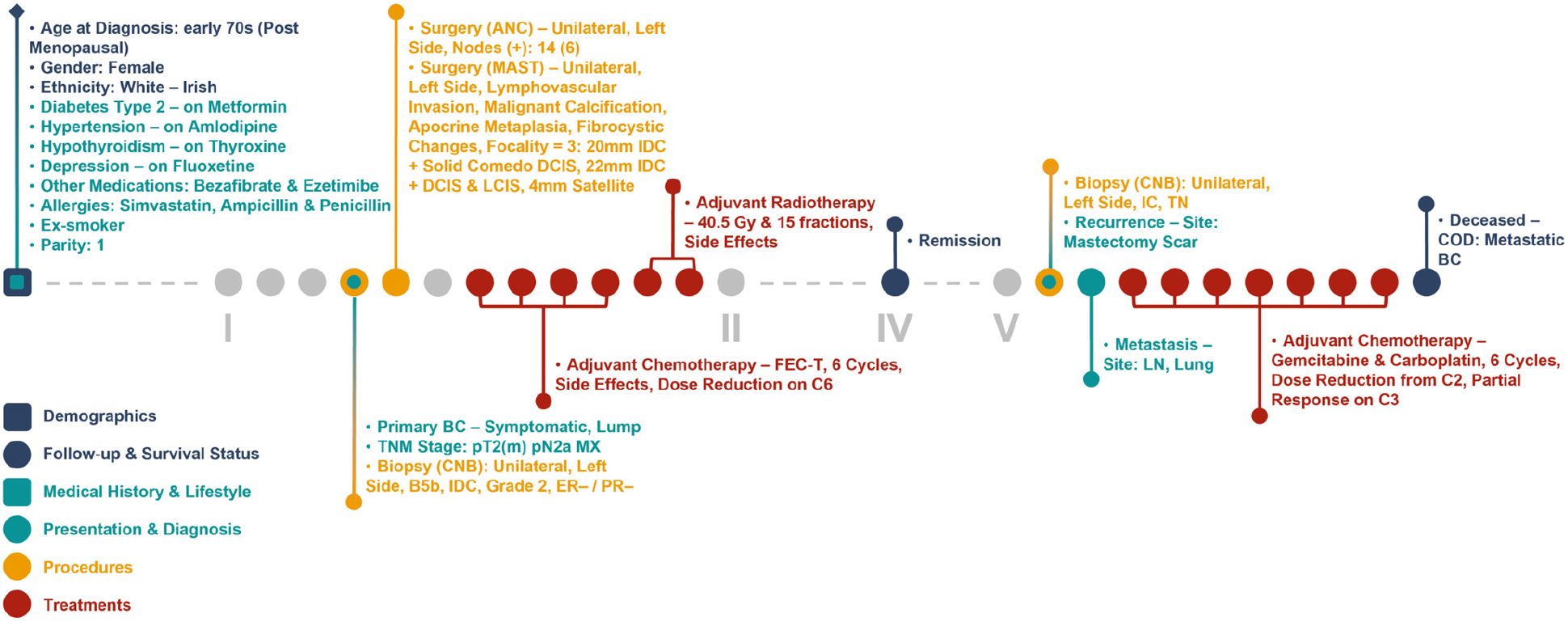
Example patient journey based on BCNTB-BCI data

Figures 6 and 7 show the clinical patient journey narratives for the same breast cancer patient based on the structured and unstructured parts (specifically imaging reports) of their EHRs from BH, respectively. EHRs differ from biobank data in terms of what information is recorded, and when and how this information is recorded: Compared to BCNTB-BCI data, EHRs from BH provide a better coverage of clinical events, which makes them suitable for certain studies (e.g., comorbidity analysis); however, they are not necessarily cancer-specific, and therefore, require filtering before they can be used to visualise cancer patient trajectories. In BCNTB-BCI data, clinical events appear sooner and with a single date, whereas in EHRs from BH, codes may have been entered multiple times with different dates. As a cancer-oriented biobank, BCNTB records cancer-related information explicitly and mostly in a structured format, while in EHRs, the same information may not be directly available or are recorded in an unstructured format. For instance, BCNTB-BCI records age at diagnosis and treatment regimen, including the response and any side effects, explicitly. In contrast, EHRs from BH calculate age at diagnosis based on date of birth and date of the earliest instance of the relevant diagnosis code; drugs that are part of the same regimen are not grouped together; and the response to treatment and any side effects are usually mentioned in free-text reports.

**Figure 6.**
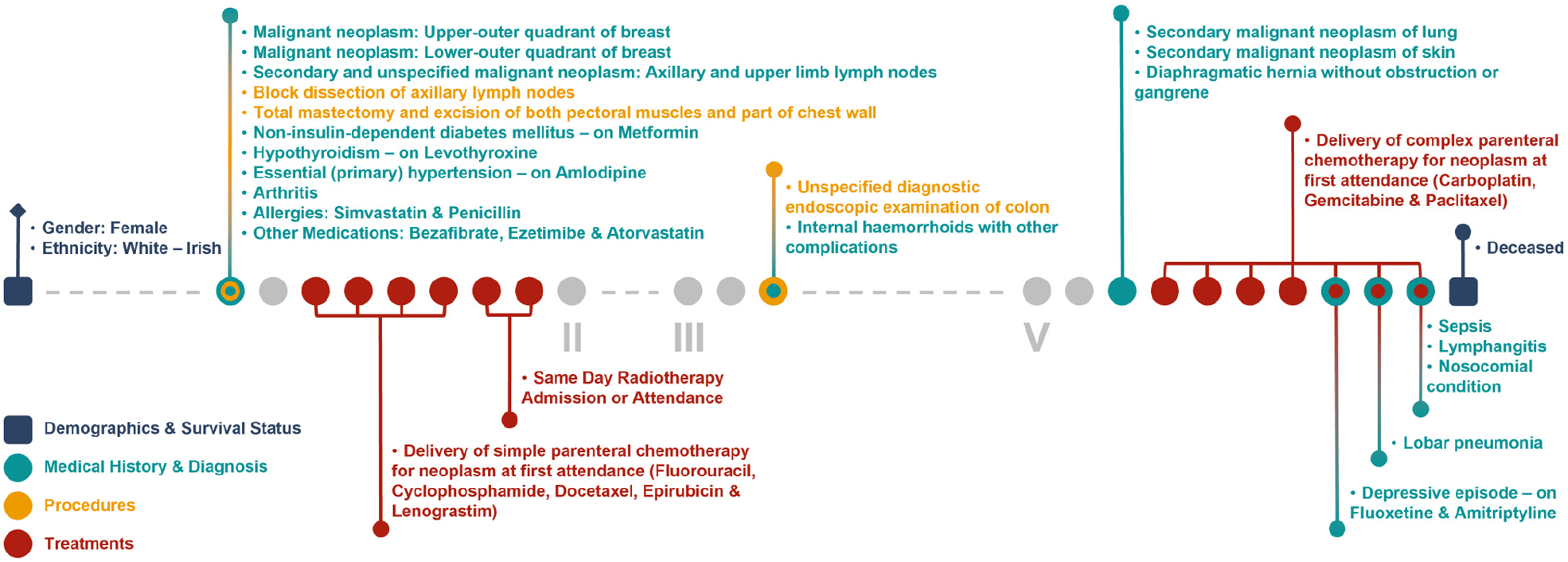
Example patient journey based on the structured parts of EHRs from BH (non-cancer-related information are omitted; ICD-10 and OPCS-4 descriptions are shown for diagnoses and procedures, respectively; drugs are shown next to the condition they were prescribed for).

**Figure 7.**
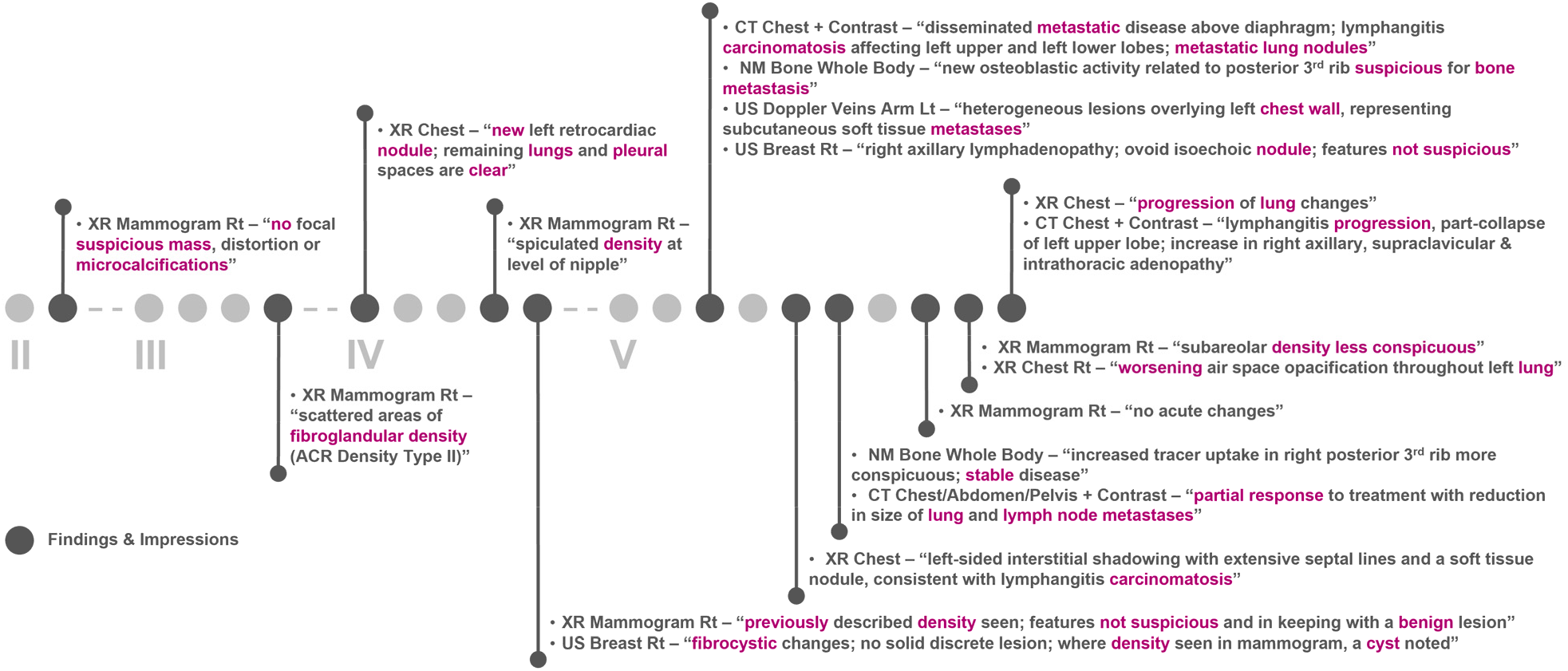
Example patient journey based on imaging reports in EHRs from BH (non-cancer-related information are omitted; keywords highlighted by the NLP algorithm are shown in purple; reports are paraphrased and summarised).

Figure 8 shows associations between gene alterations in the same breast cancer patient’s genome from 100K GP and drug response, analysed on the Analytics Hub platform using the Cancer Genome Interpreter (CGI) [28]. Genomic insights, in conjunction with information from other sources (e.g., biobank data and EHRs), not only provide researchers with richer datasets, but also guide clinicians in their diagnosis and management decision-making process. Note here the patientʹs history of having been administered Carboplatin according to their EHRs despite the patientʹs resistance to Cisplatin based on their 100K GP data analysed on Analytics Hub. The two drugs are cross-resistant [29, 30]. This is in line with the patientʹs initial partial response followed by disease progression according to their BCNTB-BCI data.

**Figure 8.**
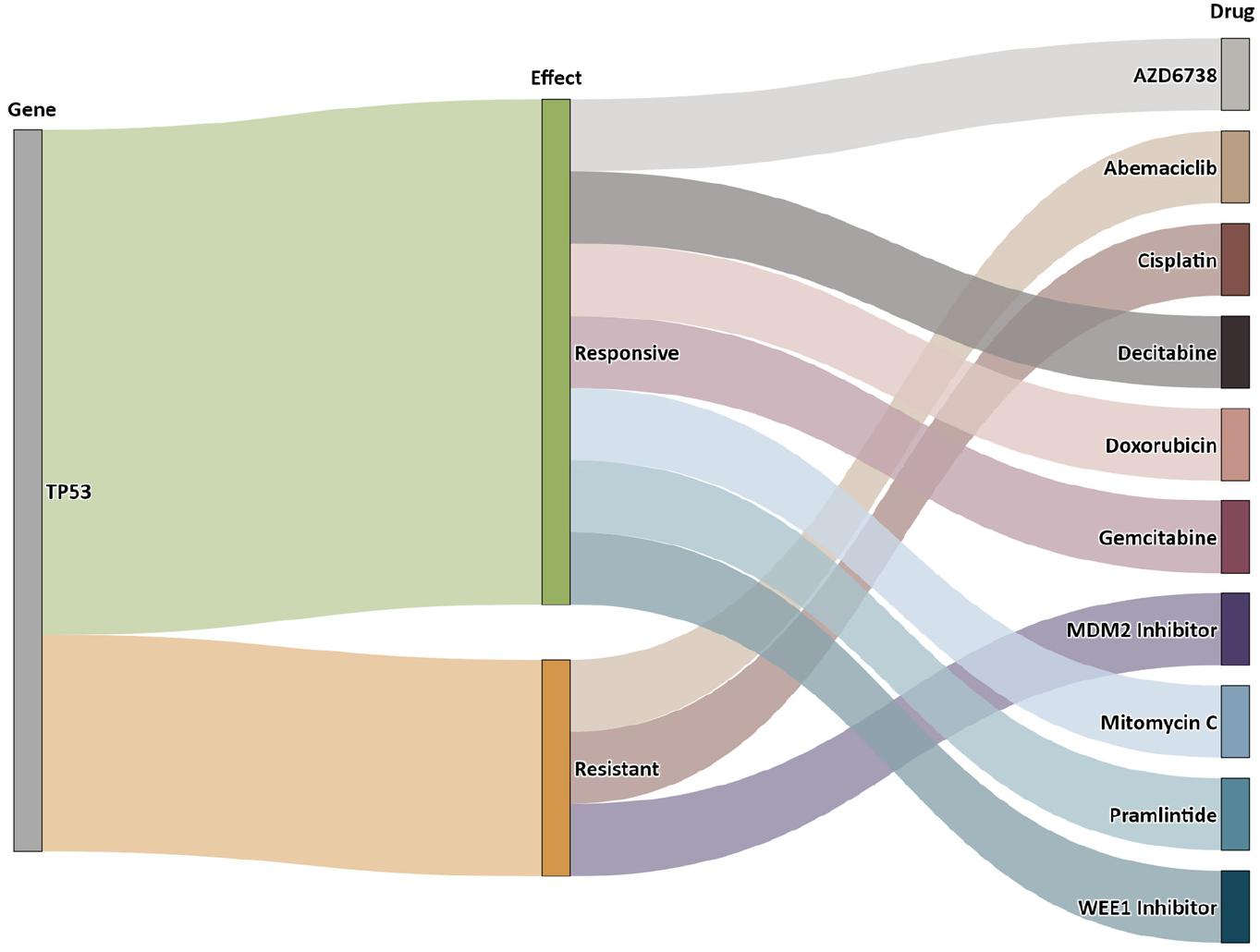
Example of a patient’s sequencing data from 100K GP analysed on Analytics Hub (only gene-drug pairs where alteration match is complete are shown).

## 4. Discussion

We adopted a dynamic infrastructure using REDCap for the management of BCNTB data and provision of its biospecimens. We implemented and customised measures that improve the quality (correctness, completeness, consistency and currency or timeliness) of the Bank’s data. Additionally, mapping our data to a standard biomedical ontology (the SNOMED-CT concepts) facilitated their integration and interoperability with EHRs and genomic data. This resulted in a rich longitudinal multimodal dataset that captures the breast cancer landscape in a multi-ethnic population and highlights the shortcomings in detection, management and treatment of breast cancer in the local area. Previous studies examining breast cancer presentation in this geographical region carried out surveys (via patient questionnaires and interviews) to collect data, which rely on large samples that are representative of the target population and high response rates [31]. BCNTB-BCI data can supplement, validate and guide such studies.

The BCNTB integrated Biospecimen and Data Request System offers real-time information on the availability of biospecimens and data to researchers, while simplifying the data sharing process to maximise data reuse. The BCNTB integrated Bioinformatics Portal enables the research community not only to access high-quality annotated biospecimens, but also to interrogate the associated data. Analysing various types of data available for each patient is key to implementing personalised diagnostic, prognostic and therapeutic strategies. It reduces research time and complexity, while the seamless data exchange and navigation between Sample Finder and Analytics Hub helps overcome the barriers due to lack of data compatibility. This created a domain-specific repository of breast cancer data and tools that can be used to generate clinical and molecular patient journey narratives. Such narratives offer context and a cross-functional view of patient experience that cannot be achieved without employing these tools and by interrogating data from either source alone, and engage all stakeholders to identify gaps in breast cancer care.

## 5. Conclusions

We present BCNTB-BCI as a dynamic cancer biobank with an infrastructure that includes several features to help tackle challenges in breast cancer research. They include: Implementation of a flexible yet robust data management system; linkage to other health data sources; and integrated data access and analytics tools. We demonstrated how these features improve data quality, increase interoperability, encourage data reuse and accelerate research. Specifically, we showcased an example where such a biobanking ecosystem can aid in predicting the health trajectory of breast cancer patients and identifying actionable opportunities for intervention.

## Data Availability

All data produced in the present study are available upon reasonable request to the authors.

## Supplementary Materials

None.

## Author Contributions

Conceptualisation, Ma.A., E.G. and C.C.; methodology, Ma.A., E.G. and C.C.; software, Ma.A., E.G., Mi.A. and J.O.; validation, R.B.; formal analysis, Ma.A. and E.G.; investigation, Ma.A. and E.G.; resources, L.J. and C.C.; data curation, Ma.A., E.G.; writing—original draft preparation, Ma.A. and E.G.; writing—review and editing, Ma.A., E.G., Mi.A., J.O., R.B., S.J., C.C. and L.J.; visualisation, Ma.A., E.G. and Mi.A.; supervision, C.C.; project administration, C.C.; funding acquisition, L.J. and C.C. All authors have read and agreed to the published version of the manuscript.

## Acknowledgments

This paper is dedicated to all the individuals who have donated to BCNTB. We are grateful to members of BCNTB-BCI (Rachel Nelan, Jennifer McGuinness, Jenny Gomm and Iain Goulding) and other BCNTB centres (led by Angela Cox and Valerie Speirs) for their help in setting up the framework for collection and distribution of biospecimens.

## References

1. Cancer Research UK. Breast cancer statistics. Available online: https://www.cancerresearchuk.org/health-professional/cancer-statistics/statistics-by-cancer-type/breast-cancer (accessed on 1 October 2022).

2. Pan, H.; Gray, R.; Braybrooke, J.; et al. 20-year risks of breast-cancer recurrence after stopping endocrine therapy at 5 years. N Engl J Med 2017, 377(19), pp. 1836–1846.

3. Breast Cancer Now. Almost one million women in UK miss vital breast screening due to COVID-19. Available online: https://breastcancernow.org/about-us/media/press-releases/almost-one-million-women-in-uk-miss-vital-breast-screening-due-covid-19 (accessed on 1 October 2022).

4. World Health Organization. The International Statistical Classification of Diseases and Health Related Problems, 10th Revision, Volume 1, Tabular List. 5th ed.; WHO: Geneva, Switzerland, 2016.

5. NHS Digital. OPCS Classification of Interventions and Procedures. Available online: https://datadictionary.nhs.uk/support-ing_information/opcs_classification_of_interventions_and_procedures.html (accessed on 1 October 2022).

6. National Institutes of Health. Systematized Nomenclature of Medicine – Clinical Terms. Available online: https://www.nlm.nih.gov/healthit/snomedct (accessed on 1 October 2022).

7. Breast Cancer Now. Breast Cancer Now Tissue Bank. Available online: https://breastcancernow.org/breast-cancer-re-search/breast-cancer-now-tissue-bank (accessed on 1 October 2022).

8. Felmeister, A.S.; Rivera, T.J.; Masino, A.J.; et al. Scalable biobanking: a modular electronic honest broker and biorepository for integrated clinical, specimen and genomic research. In Proceedings of the IEEE International Conference on Bioinformatics and Biomedicine (BIBM), Washington, D.C., USA, 2015; pp. 484–490.

9. Kinkorová, J. Biobanks in the era of personalized medicine: objectives, challenges, and innovation. EPMA J 2016, 7(1), pp. 1–12.

10. Olson, J.E.; Bielinski, S.J.; Ryu, E.; et al. Biobanks and personalized medicine. Clin Genet 2014, 86(1), pp. 50–55.

11. NHS North East London Clinical Commissioning Group. East London Patient Record. Available online: https://northeastlon-donccg.nhs.uk/about-us/elpr.htm (accessed on 1 July 2021).

12. Harris, P.A.; Taylor, R.; Thielke, R.; et al. Research electronic data capture (REDCap) – a metadata-driven methodology and workflow process for providing translational research informatics support. J Biomed Inform 2009, 42(2), pp. 377–381.

13. Harris, P.A.; Taylor, R.; Minor, B.L.; et al. The REDCap consortium: Building an international community of software platform partners. J Biomed Inform 2019, 95, p. 103208.

14. Barts Health NHS Trust. Fast facts. Available online: https://www.bartshealth.nhs.uk/fast-facts (accessed on 1 October 2022).

15. Somerset NHS Foundation Trust. Somerset Cancer Register. Available online: https://www.somersetft.nhs.uk/somerset-cancer-register (accessed on 1 October 2022).

16. NHS Digital. Data collections and data sets. Available online: https://digital.nhs.uk/data-and-information/data-collections-and-data-sets (accessed on 1 October 2022).

17. Centre for Genomic Regulation and European Bioinformatics Institute. European Genome-phenome Archive. Available online: https://ega-archive.org (accessed on 1 October 2022).

18. 1000 Genomes Project Consortium. A global reference for human genetic variation. Nature 2015, 526(7571), p. 68.

19. The National Cancer Intelligence Network. The National Cancer Registration and Analysis Service. Available online: http://www.ncin.org.uk (accessed on 1 October 2022).

20. Grinberg, M. Flask web development: developing web applications with python. OʹReilly Media, Inc., 2018.

21. Gentleman, R.C.; Carey, V.J.; Bates, D.M.; et al. Bioconductor: open software development for computational biology and bioinformatics. Genome Biol 2004, 5(10), pp. 1–16.

22. Oscanoa, J.; Sivapalan, L.; Gadaleta, E.; et al. SNPnexus: a web server for functional annotation of human genome sequence variation (2020 update). Nucleic Acids Res 2020, 48(W1), pp. W185–192.

23. Gadaleta, E.; Fourgoux, P.; Pirró S.; et al. Characterization of four subtypes in morphologically normal tissue excised proximal and distal to breast cancer. NPJ Breast Cancer 2020, 6(1), pp. 1–12.

24. National Cancer Institute. The Cancer Genome Atlas. Available online: https://www.cancer.gov/about-nci/organization/ccg/re-search/structural-genomics/tcga (accessed on 1 October 2022).

25. The Broad Institute of Harvard and MIT. Cancer Cell Line Encyclopedia. Available online: https://sites.broadinstitute.org/ccle (accessed on 1 October 2022).

26. Ng, H.S.; Vitry, A.; Koczwara, B.; et al. Patterns of comorbidities in women with breast cancer: a Canadian population-based study. Cancer Causes Control 2019, 30(9), pp. 931–941.

27. Elixhauser, A.; Steiner, C.; Harris, D.R.; et al. Comorbidity measures for use with administrative data. Med Care 1998, pp. 8–27.

28. BBGLab. Cancer Genome Interpreter. Available online: https://www.cancergenomeinterpreter.org (accessed on 1 October 2022).

29. Rabik, C.A.; Dolan, M.E. Molecular mechanisms of resistance and toxicity associated with platinating agents. Cancer Treat Rev 2007, 33(1), pp. 9–23.

30. Galluzzi, L.; Senovilla, L.; Vitale, I.; et al. Molecular mechanisms of cisplatin resistance. Oncogene 2012, 31(15), pp. 1869–1883.

31. Forbes, L.J.L.; Atkins, L.; Thurnham, A.; et al. Breast cancer awareness and barriers to symptomatic presentation among women from different ethnic groups in East London. Br J Cancer 2011, 105(10), pp. 1474–1479.

